## Supplementary material for "Fat tails and the need to disclose distribution parameters of qEEG databases": Testing more fat-tailed distributions

**Reproduction of results using several fat-tailed distributions**

To demonstrate the generality of our results we also calculated the excess of false-positives using other three fat-tailed distributions, the t-distribution with a low number of degrees of freedom, the Chi²-distribution also with a low number of degrees of freedom and Pareto distributions with different exponents. Results showed in the Figures S1 to S3, below, clearly show that the inflation in the number of false-positives is common to all fat-tailed distributions.

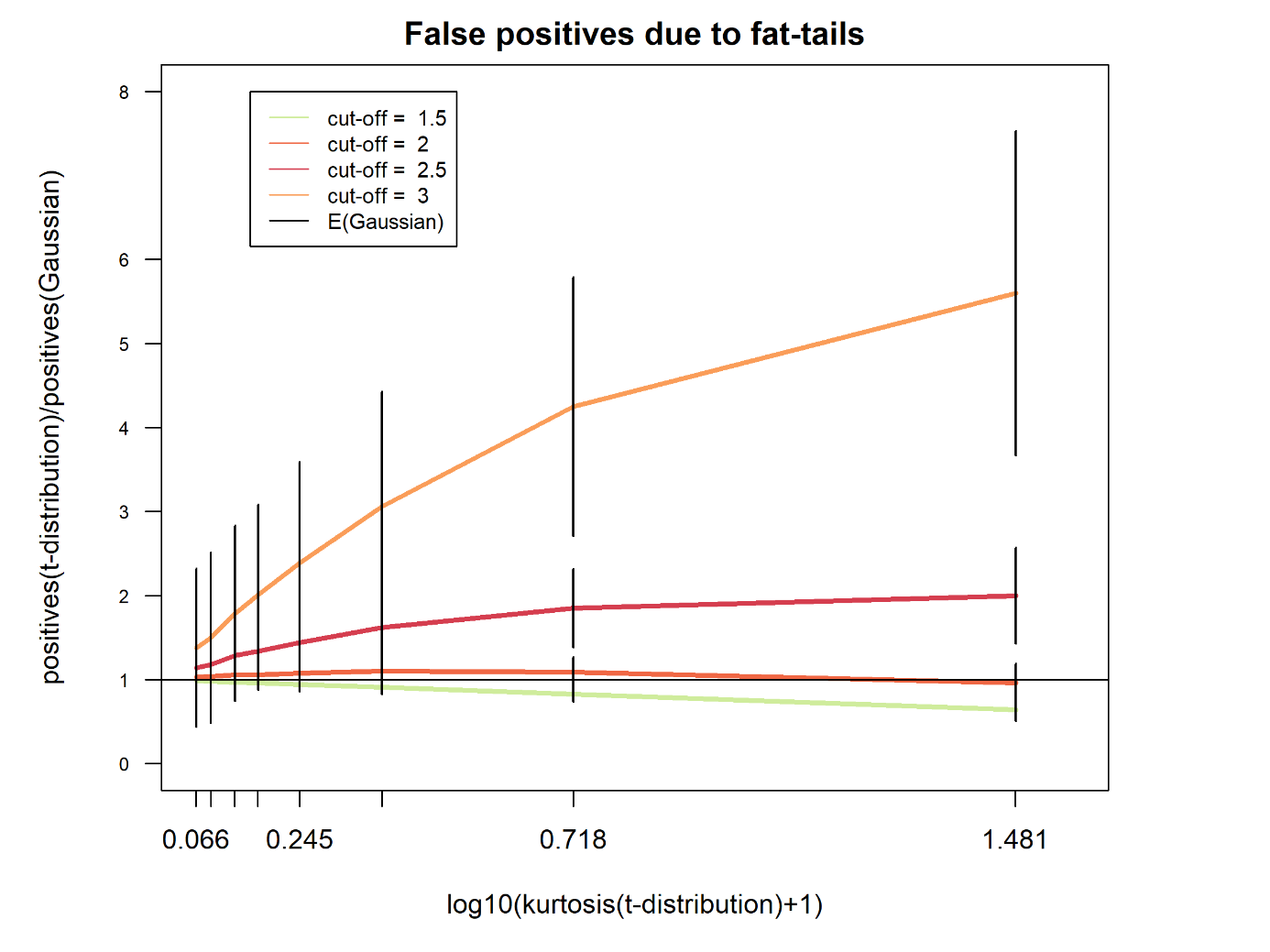

t-distribution: We generated samples of n=1000 observations using the t-distribution with 3, 5, 8, 12, 16, 20, 30 and 40 degrees-of-freedom. The lower the degrees of freedom of a t-distribution, the more fat-tailed it is. In the Figure S1 the ratio of false-positives(t-distribution)/false-positives(Gaussian) is displayed as a function of the ratio kurtosis(t-distribution)/kurtosis(Gaussian). The larger this last ratio, the more fat-tailed is the distribution.

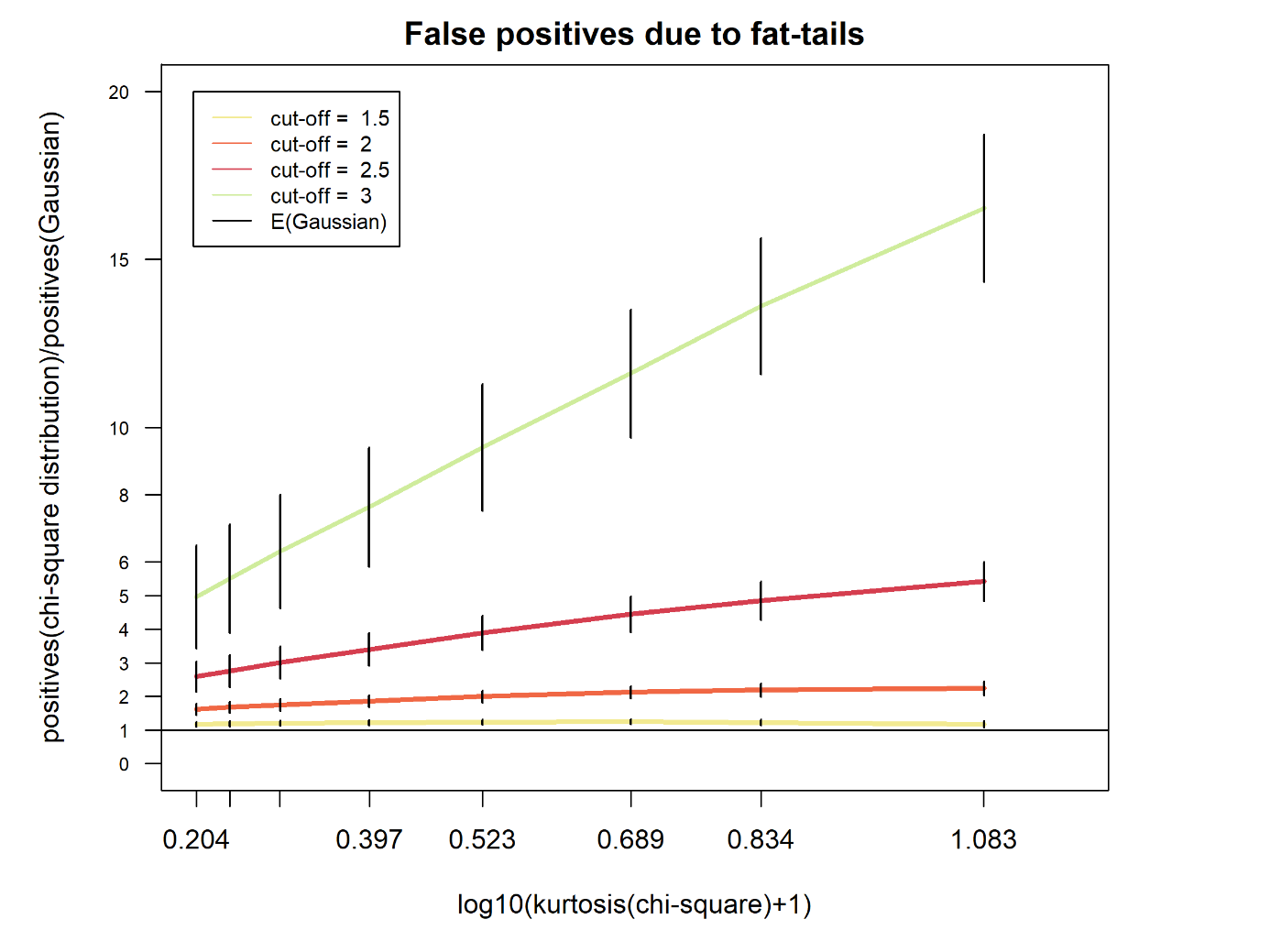

Chi²-distribution: We generated samples of n=1000 observations using the chi²-distribution with 1, 2, 3, 5, 8, 12, 16, and 20 degrees-of-freedom. The lower the degrees of freedom of a chi²-distribution, the more fat-tailed it is (Fleishman, 1978, p. 528). In the Figure S2 the ratio of false-positives(chi²-distribution)/false-positives(Gaussian) is displayed as a function of the ratio kurtosis(chi²-distribution)/kurtosis(Gaussian). The larger this last ratio, the more fat-tailed is the distribution.

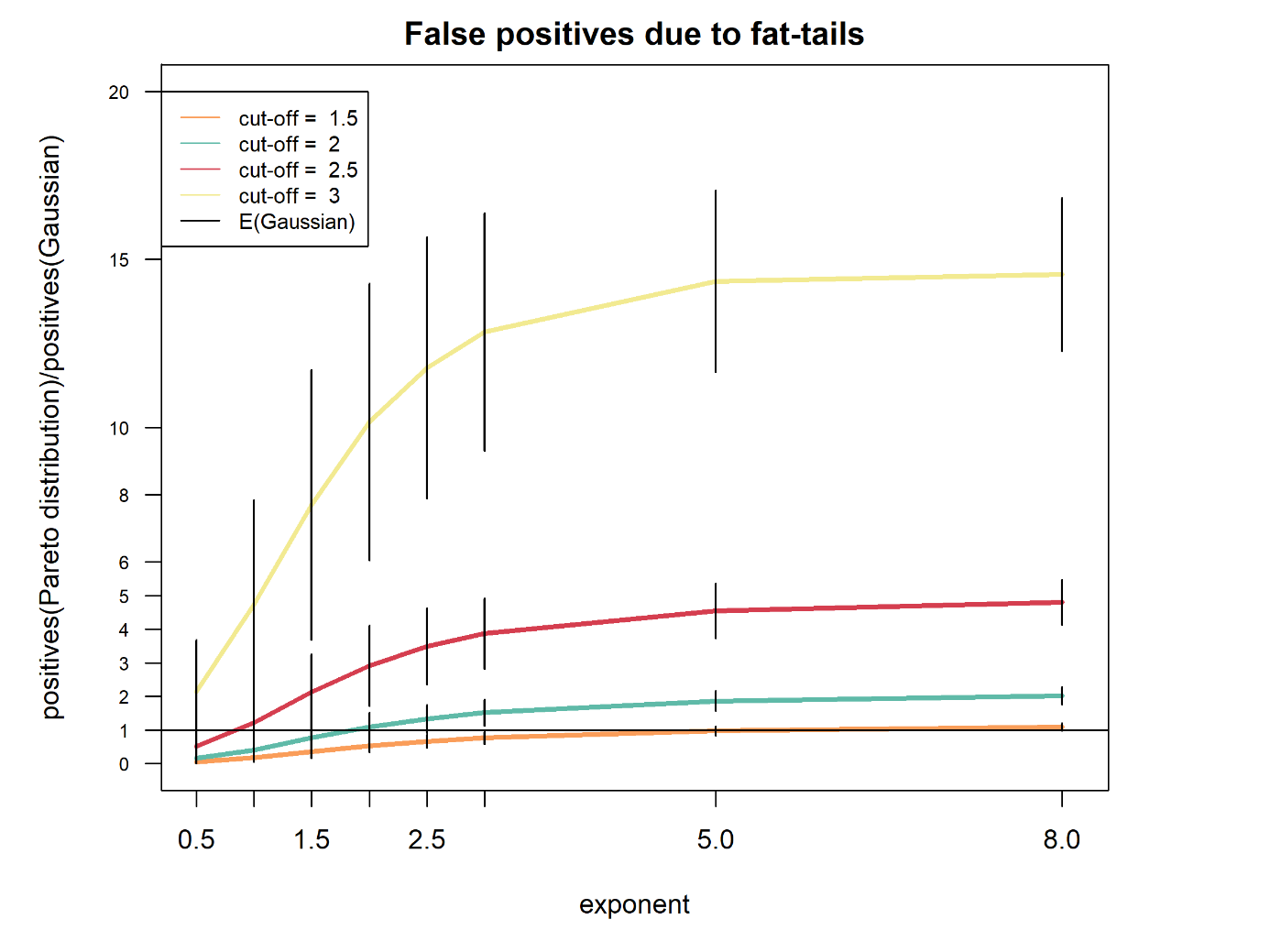
Pareto distribution: We generated samples of n=1000 observations using the Pareto distribution with a location of 1 and exponents 0.5, 1, 1.5, 2, 2.5, 3, 5, and 8. The lower the exponent of a Pareto distribution, the more fat-tailed it is. In the Figure S3 the ratio of false-positives(Pareto distribution)/false-positives(Gaussian) is displayed as a function of the exponent of the distribution. The difference in the x-axis to the other plots is due to the fact that the mean, variance, and other moments are finite only if the shape parameter a is sufficiently large. The larger this last ratio, the more fat-tailed is the distribution.

**References**

Fleishman, A. I. (1978). A method for simulating non-normal distributions. *Psychometrika*, *43*(4), 521-532.
